# Mitigation of gastrointestinal graft versus host disease with tocilizumab prophylaxis is accompanied by preservation of microbial diversity and attenuation of enterococcal domination

**DOI:** 10.1101/2022.06.30.22277104

**Authors:** Saurabh Chhabra, Aniko Szabo, Annelie Clurman, Katelynn McShane, Nicholas Waters, Daniel Eastwood, Lisa Samanas, Teng Fei, Gabriel Armijo, Sameen Abedin, Walter Longo, Parameswaran Hari, Mehdi Hamadani, Nirav N. Shah, Lyndsey Runaas, James H. Jerkins, Marcel van den Brink, Jonathan U. Peled, William R. Drobyski

## Abstract

A common feature in the gastrointestinal (GI) tract during allogeneic hematopoietic stem cell transplantation is the loss of microbial diversity and emergence of opportunistic pathogens that can adversely impact survival. Consequently, preventing transplant-associated dysbiosis is an emerging strategy for optimizing treatment outcomes. In this study, we examined the effect of an extended tocilizumab administration schedule in addition to tacrolimus/methotrexate (Tac/MTX) as graft versus host disease (GVHD) prophylaxis on microbial composition in the GI tract along with overall transplant outcomes. Twenty-nine patients received busulfan-based myeloablative conditioning and were transplanted with HLA-matched related or unrelated peripheral blood stem cell grafts. The primary end point of the trial was GVHD-free relapse-free survival (GRFS) at 12 months. The cumulative incidences of grades 2-4 and 3-4 acute GVHD were 10.5% and 7% at day 180, respectively. There was one case of GVHD of the lower GI tract within the first 12 months. Non-relapse mortality and relapse-free survival were 3.4% and 86.2% at one year, respectively. GRFS was 38% at one year which was significantly higher than the pre-specified historical control rate of 20% (p=0.02) and therefore met the primary end point of the trial. Fecal samples from this patient population were sequenced and computationally analyzed centrally along with a demographically matched control cohort that received only Tac/MTX for GVHD prophylaxis. This comparative analysis revealed significantly less loss of α-diversity and reduced emergence of pathogenic organisms such as enterococcus in tocilizumab-treated recipients, demonstrating that loss of microbial diversity and enterococcal domination is attenuated in these patients. (Clinicaltrial.gov Identifier: NCT03699631).

## INTRODUCTION

Graft-versus-host disease (GVHD) is the most significant complication that occurs after allogeneic hematopoietic stem cell transplantation (HSCT). Whereas GVHD has pleiotropic clinical manifestations, the gastrointestinal (GI) tract is of primary importance during the acute phase of the disease and a major driver of morbidity and mortality.^1,2^ A characteristic observation in the GI tract in allogeneic HSCT recipients is the profound and reproducible alterations in host microbiota with an attendant loss of diversity and shift from anaerobic to more facultative aerobes.^3,4^ Dysbiosis in the microbiome has similarly been described in mice with concurrent GVHD,^5^ highlighting the role that GVHD-induced inflammation has in promoting these microbial shifts. Enrichment of specific bacteria, such as *Enterococcus*, is a characteristic feature of transplant-associated dysbiosis that predisposes patients to subsequent bacteremia and GVHD.^6-10^ Patterns of microbial disruption have been shown to be consistent across different transplant centers and geographic locations, indicative of a stereotypical microbial community response during transplantation even across different patterns of antibiotic use.^11^ Therefore, strategies to maintain diversity and remodel the intestinal microbiome offer the potential to attenuate the severity of this disease^12^ and promote tissue tolerance due to the production of commensal-derived metabolites that support epithelial integrity.^13-15^ While fecal microbiota transplantation can augment microbial diversity, this approach has been performed only in patients with either ongoing intestinal GVHD^16^ or after engraftment when dysbiosis has already been documented.^17-18^ To date, however, there have been no clinical strategies that have proven effective at preventing the development of dysbiosis in allogeneic HSCT recipients.

Interleukin 6 (IL-6) has been shown to play a pivotal role in the pathophysiology of GVHD, and blockade of this signaling pathway in pre-clinical models and clinical studies has been associated with a reduction in GVHD severity in the GI tract.^19-21^ The mechanisms by which IL-6 inhibition attenuates GVHD in pre-clinical studies includes augmentation of regulatory T cell reconstitution,^19^ a decrease in the expansion of effector T cells,^19,20,22^ and prevention of IL-6 production by dendritic cells.^22^ Whether blockade of IL-6 signaling has any salutary effect on the microbiome as another potential mechanistic pathway by which GVHD is mitigated within the GI tract has not been examined in either pre-clinical or clinical settings. To address this question, we conducted a phase 2 study in which tocilizumab was administered as adjunctive GVHD prophylaxis to patients at high risk for the development of GI tract disease to assess the effect on microbial diversity and composition, as well as on overall transplant outcomes.

## MATERIAL AND METHODS

### Patient Population

Patients were eligible if they were >18 years of age and had a diagnosis of acute leukemia, chronic myelogenous leukemia, myeloproliferative disease, or myelodysplasia with <5% marrow; had a 10/10 matched sibling or 8/8 matched unrelated donor available; ejection fraction at rest >45%; estimated creatinine clearance greater than 40 mL/minute; adjusted DLCO ≥40% and FEV1 ≥50%; and total bilirubin < 3.0x and ALT/AST <5x the upper normal limit. This protocol was approved by the Institutional Review Board of the Medical College of Wisconsin.

### Conditioning Regimens and GVHD Prophylaxis

All patients received busulfan as part of the preparative regimen. Patients receiving myeloablative conditioning (MAC)^23^ were treated with either busulfan 3.2 mg/kg/day (days -7 to -4) and cyclophosphamide 60 mg/kg/day (days -3 and -2) or busulfan 3.2 mg/kg/day (days 5- to -2) and fludarabine 30 mg/m^2^/day (days -5 to -2) (Flu/BU4). Some patients with myelofibrosis (n=3) received only three days of Busulfan (Flu/BU3). Patients all received granulocyte colony factor-stimulated peripheral blood stem cell grafts. For GVHD prevention, tacrolimus was administered intravenously at a dose of 0.03 mg/kg/day starting day –3 to maintain a level of 5-15 ng/mL. Methotrexate was given at the doses of 15 mg/m^2^ IV on day +1, and 10 mg/m^2^ IV on days +3, +6 and +11. Tocilizumab was infused intravenously at a dose of 8 mg/kg (maximum dose of 800 mg) once on day-1 approximately 24 hours prior to the hematopoietic stem cell infusion, and then again on day +100 (+/-14 days, i.e., +86 to +114) post-transplantation.

### Study Design

The primary endpoint of the study was the probability of GVHD-free/relapse-free survival (GRFS)^24^ defined as survival without grade III-IV acute GVHD, systemic therapy-requiring chronic GVHD, relapse, or death at 12 months. Pre-specified secondary objectives of the study were to define neutrophil and platelet engraftment, grades 2-4 and 3-4 acute GVHD at days 100 and 180, chronic GVHD (overall and severe), transplant related mortality (TRM), disease relapse, disease-free survival (DFS), overall survival (OS), characterization of infections, and gut microbiome diversity and composition. **Supplemental Table 1** contains the definition of the events, censorings, and competing risks for all time-to-event outcomes. The sample size was selected to achieve 80% power to detect an increase in GRFS at 12 months to 40% compared to a historical control value of 20% at a one-sided 5% significance level. Based on an asymptotic z-test for proportions, 29 patients with a known outcome were needed (i.e., known GVHD, relapse, or death status at 12 months).

### Outcome Assessments

Neutrophil recovery was defined as the first of three consecutive days with an ANC>500/mm3. Platelet engraftment was defined as the first day of a sustained platelet count above 20,000/mm3 without any platelet transfusions for the preceding seven days. The grade of acute GVHD was determined using the Harris criteria.^25^ Chronic GVHD was graded using the NIH Chronic GVHD Consensus criteria.^26^ GVHD events were adjudicated by an independent review panel (IRP) and the protocol investigators were blinded (or masked) to the IRP and were not permitted to grade or modify GVHD assessments.

### Other detailed methods

All other additional methodology related to the clinical trial, microbiome sequencing, and statistical analyses of clinical outcomes and microbiome comparisons are described in Supplemental Methods.

## RESULTS

### Patient Characteristics

Between November 21, 2018, and February 12, 2020, 29 patients were enrolled in the study. The demographic data for this population is detailed in **Table 1**. The median age of the cohort was 57 (range 24-66). All patients received Busulfan (BU)-based conditioning. Twenty-five patients received Flu/BU4, three were treated with Flu/BU3 and one received Bu/CY for myeloablative conditioning. All patients received peripheral blood stem cell grafts from related (n=12) or matched unrelated (n=17) donors. Diseases consisted of de novo AML (n=14), secondary AML (n=1), ALL (n=1), MDS (n=6), Myelofibrosis (n=5), and CML (n=2). Using the adjusted disease risk index,^27^ patients were classified as intermediate (n=12), high (n=16), or very high (n=1) risk. The HCT-CI scores were 0-1 (n=10), 2 (n=7), 3 (n=6), and 4-5 (n=6) in the patient cohort. The median Karnofsky score was 80 (range 70-100). All patients received a first dose of tocilizumab on day -1. Twenty-six patients received a second dose a median of 99 days post transplantation (range 86-114 days). Three patients did not receive a second dose; one due to relapse prior to day 100, one developed grade 3-4 acute GVHD which triggered the GRFS endpoint, and the last patient refused due to concerns about acquiring SARS-CoV2 infection during outpatient tocilizumab administration. Median follow up for surviving patients was 20.6 months (range 14.4-29.2).

**TABLE 1.**
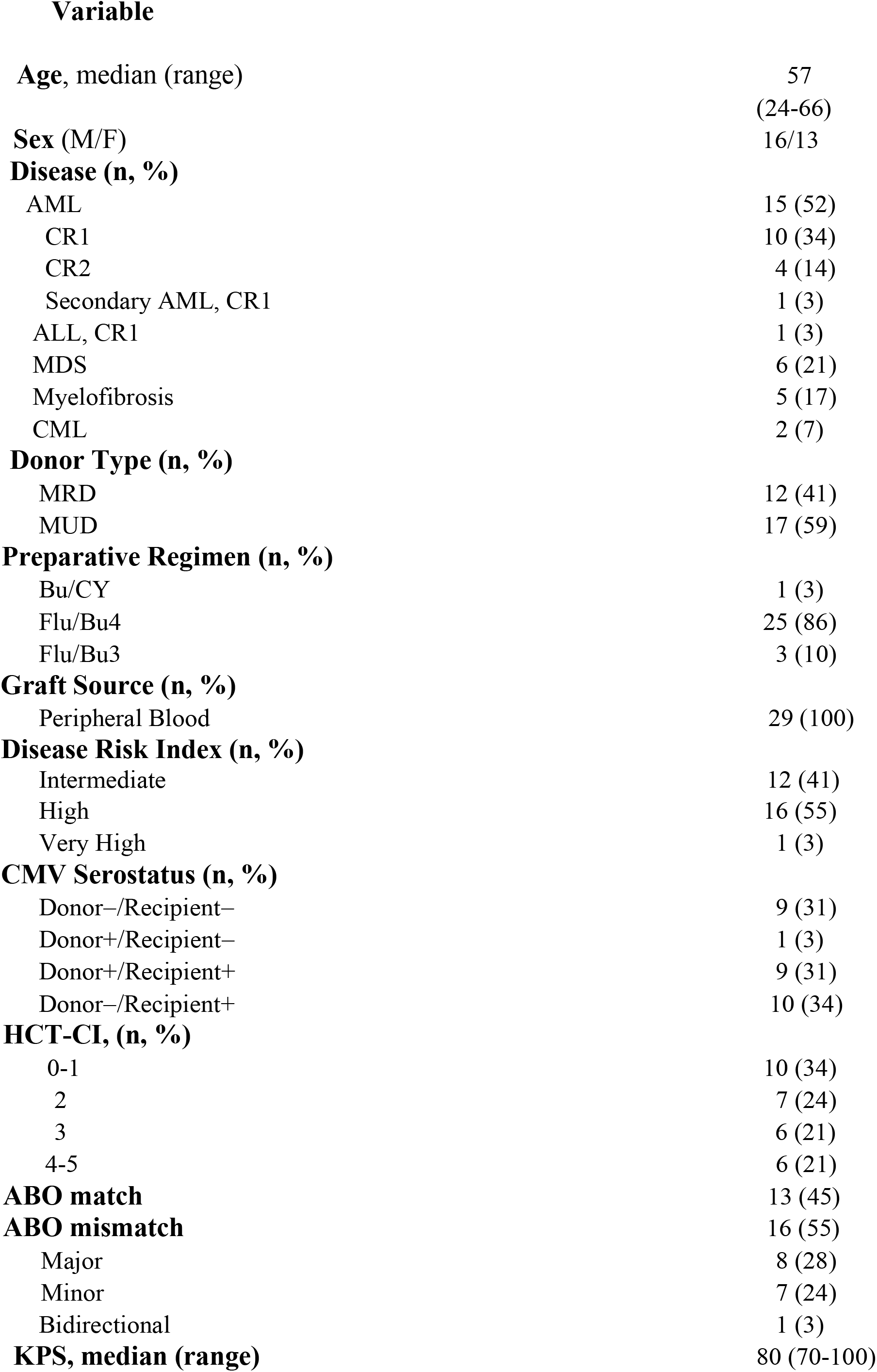
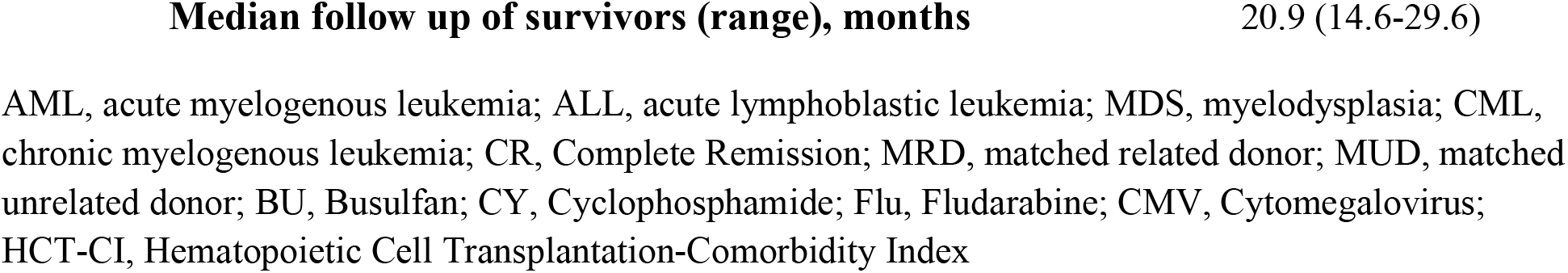
PATIENT CHARACTERISTICS OF TOCILIZUMAB TRIAL PATIENTS (N=29)

### Engraftment and Chimerism

There were no cases of primary graft failure. The median time to engraftment was 16 days (range 13-26) for neutrophils and 19 days (range 10-35) for platelets (**Supplemental Figure 1**). Chimerism studies were conducted on days 30 and 100 post transplantation in all patients. Median donor CD3 chimerism was 89% (range 66-100), and 92% (range 73-100) on days 30 and 100, respectively, while median CD33 chimerism was 100% (range 99-100), and 100% (range 99-100) at the same time points.

### GVHD, Transplant-related Mortality, Relapse, Survival, and GRFS

The cumulative incidence of grades 2-4 acute GVHD at day 180 was 10.5% (95% CI 2.6-24.9%) (**Figure 1A**), whereas the incidence of grades 3-4 acute GVHD was 7.0% (95% CI 1.2-20.4) (**Figure 1B**). One patient had grade 2 acute GVHD due to stage 3 skin involvement. Two patients developed grade 3 acute GVHD; one that involved the lower GI tract and another with stage 2 liver and biopsy-proven upper GI involvement. There were no other cases of lower GI tract involvement within the first 12 months. Four of 29 patients developed bacteremia within the first 100 days (i.e., *Escherichia coli, Capnocytophaga sputigena, Streptococcus oralis, and Enterococcus faecalis*). These blood stream infections occurred on days +10, +11, +15 and +48 post transplantation, respectively. The cumulative incidence of overall chronic GVHD was 64.8% (95% CI 42.4-80.3) (**Figure 1C**), whereas the incidence of severe chronic GVHD^26^ was 7.7% (95% CI 1.3-22.1) at 12 months (**Figure 1D**). Organ involvement in the 17 patients who developed chronic GVHD consisted of liver (10), mouth (7), eyes (5), skin (4), lung (1), genital (1), and joints (1) (**Table 2**). The cumulative incidence of relapse was 10.3% (95% CI 2.5-24.6) at one year (**Figure 1E**). Non-relapse mortality was 3.4% (95% CI 0.2-15.3) at 12 months and occurred in one patient due to bronchiolitis obliterans in the setting of pre-existing interstitial lung disease prior to transplantation (**Figure 1F**). Overall survival and relapse-free survival were identical at 12 months (86.2%; 95% CI 67.3-94.6) (**Figure 1G and 1H**). The probability of GRFS at one year was 37.9% (95% CI 20.9-54.9) which was significantly higher than the pre-specified historical rate of 20% (p = 0.023), indicating that the primary end point of the trial was met (**Figure 1I**). The median time to triggering GRFS was 8.5 months.

**Figure 1:**
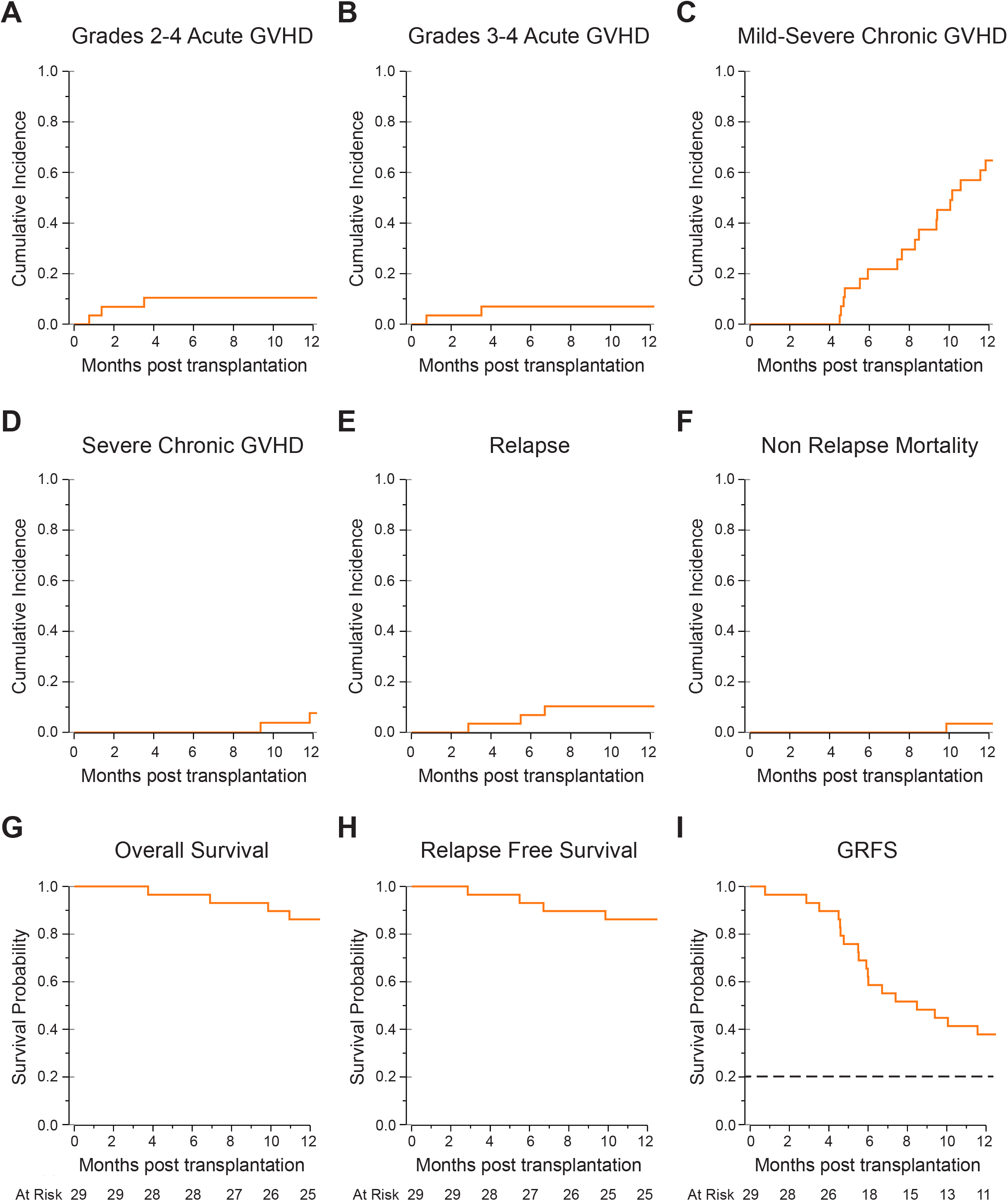
Transplant Outcomes in Patients that received GVHD Prophylaxis with Tocilizumab. (A, B). Cumulative incidences of grades 2-4 (panel A) and 3-4 (panel B) acute GVHD. (C, D). Cumulative incidences of mild-severe and severe chronic GVHD. (E). Cumulative incidence of relapse. (F). Cumulative incidence of non-relapse mortality. (G, H). Probability of overall survival (panel G) and relapse-free survival (panel H). (I). Probability of GVHD-free/relapse-free survival (GRFS). Dotted line denotes the pre-specified historical control rate of 20%.

**TABLE 2.** SEVERITY OF CHRONIC GVHD BY NIH GRADE.

| **Disease Severity (n)** | **Organs Involved (n)** |
| --- | --- |
| Mild Chronic GVHD (8) | Eye (3)<br>Mouth (3)<br>Liver (4)<br>Skin (2)<br>Eosinophilia (1)<br>Joint Swelling (1) |
| Moderate Chronic GVHD (7) | Eyes (1)<br>Mouth (3)<br>Liver (5)<br>Skin (1)<br>GU (1) |
| Severe Chronic GVHD (3) | Eyes (1)<br>Mouth (1)<br>Skin (2)<br>Liver (1)<br>Lungs (1)<br>Eosinophilia (1) |

The maximum severity of chronic GVHD at one year post transplantation is shown along with the organs involved in each disease severity category.

### Comparative Microbiome Analysis

To examine the fecal microbial composition in patients that received GVHD prophylaxis with tocilizumab, we compared evaluable stool samples from trial participants (toci) with those collected from an observational cohort at Memorial Sloan Kettering Cancer Center (MSKCC) (control). Patients in this comparison were required to have an evaluable baseline stool sample collected between Day-7 and Day-1 relative to transplant. Fecal samples were obtained from MCW trial participants prior to transplantation, and then weekly through the period of engraftment. A total of 122 samples were collected from 27 patients that received tocilizumab. The first two patients in the trial were not included since the amendment which allowed us to collect, process and ship fecal samples had not been approved by the MCW IRB until after the first two patients had been enrolled. Of the 27 sampled recipients, 1 patient did not provide a sample prior to transplantation and an additional 7 baseline samples were not evaluable for reasons of amplification or sequencing failure. Thus, 19 recipients had evaluable baseline samples and, in aggregate, there were 82 evaluable samples from these patients who comprised the “toci” cohort. The control population was assembled from demographically matched patients at MSKCC who contributed to a fecal biobank and had received an unmodified peripheral blood stem cell transplant following myeloablative conditioning with Flu/BU4 and received tacrolimus and methotrexate as sole GVHD prophylaxis (**Table 3)**. There were 38 patients who met these criteria and also had a baseline stool sample collected between Day-7 and Day-1 prior to transplantation; this comprised the microbiome control cohort which consisted of 308 total samples.

**TABLE 3.**
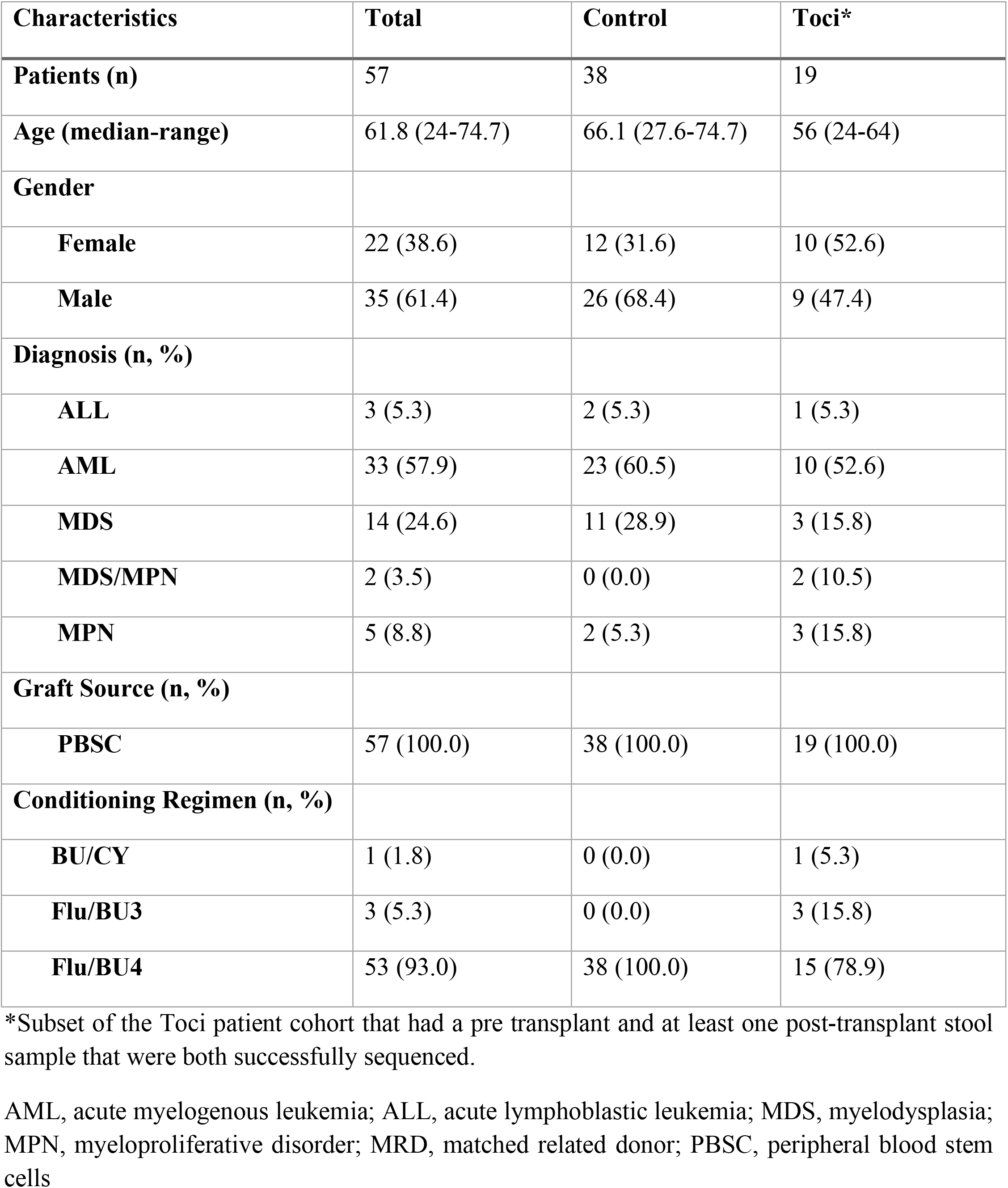
DEMOGRAPHIC CHARACTERISTICS OF THE TOCILIZUMAB SUBSET AND THE MSK CONTROL COHORT FOR MICROBIOME ANALYSIS.

| **Characteristics** | **Total** | **Control** | **Toci\*** |
| --- | --- | --- | --- |
| **Patients (n)** | 57 | 38 | 19 |
| **Age (median-range)** | 61.8 (24-74.7) | 66.1 (27.6-74.7) | 56 (24-64) |
| **Gender** |  |  |  |
| **Female** | 22 (38.6) | 12 (31.6) | 10 (52.6) |
| **Male** | 35 (61.4) | 26 (68.4) | 9 (47.4) |
| **Diagnosis (n, %)** |  |  |  |
| **ALL** | 3 (5.3) | 2 (5.3) | 1 (5.3) |
| **AML** | 33 (57.9) | 23 (60.5) | 10 (52.6) |
| **MDS** | 14 (24.6) | 11 (28.9) | 3 (15.8) |
| **MDS/MPN** | 2 (3.5) | 0 (0.0) | 2 (10.5) |
| **MPN** | 5 (8.8) | 2 (5.3) | 3 (15.8) |
| **Graft Source (n, %)** |  |  |  |
| **PBSC** | 57 (100.0) | 38 (100.0) | 19 (100.0) |
| **Conditioning Regimen (n, %)** |  |  |  |
| **BU/CY** | 1 (1.8) | 0 (0.0) | 1 (5.3) |
| **Flu/BU3** | 3 (5.3) | 0 (0.0) | 3 (15.8) |
| **Flu/BU4** | 53 (93.0) | 38 (100.0) | 15 (78.9) |

\*Subset of the Toci patient cohort that had a pre transplant and at least one post-transplant stool sample that were both successfully sequenced.
AML, acute myelogenous leukemia; ALL, acute lymphoblastic leukemia; MDS, myelodysplasia; MPN, myeloproliferative disorder; MRD, matched related donor; PBSC, peripheral blood stem cells

A comparison of baseline microbiomes demonstrated that α-diversity was significantly lower in toci patients prior to transplantation (**Figure 2A**). We next examined the relative baseline abundances of bacteria assigned to four genera of interest (*Blautia, Enterococcus, Eschericia*, and *Akkermansia*) in allogeneic HSCT patients. These genera were selected based on prior studies which have described associations for *Blautia* with lower rates of GVHD-related mortality,^28^ *Enterococcus* as a pathogen in GVHD in humans and murine recipients,^6,10,29^ *Eschericia* as a cause of bloodstream infections in patients,^7,30^ and *Akkermansia* as an emergent bacterium in mouse models of GVHD.^31^ In this analysis, we observed that the toci cohort had a lower abundance of members of genus *Blautia* and *Enterococcus*, whereas there was a higher abundance of *Akkermansia* and *Eschericia* (**Figure 2B**). Principal coordinate analysis (PCoA) of global composition, in which each point is a sample and samples with similar composition are closer together, revealed that variation along the first coordinate was attributable primarily to the control cohort (**Figure 2C**). Subsetting this PCoA plot by time intervals demonstrated that at baseline, samples from both the toci and control cohorts clustered at lower values of PCo1. A shift to the right of the plot, (i.e., higher values of PCo1) were observed over time more prominently among control (blue) than toci (red) samples, particularly at days 7 and 14 (**Figure 2D**). This indicated that the samples from the tocilizumab cohort were spared the perturbation in microbiome composition that is summarized by the first principal coordinate.

**Figure 2:**
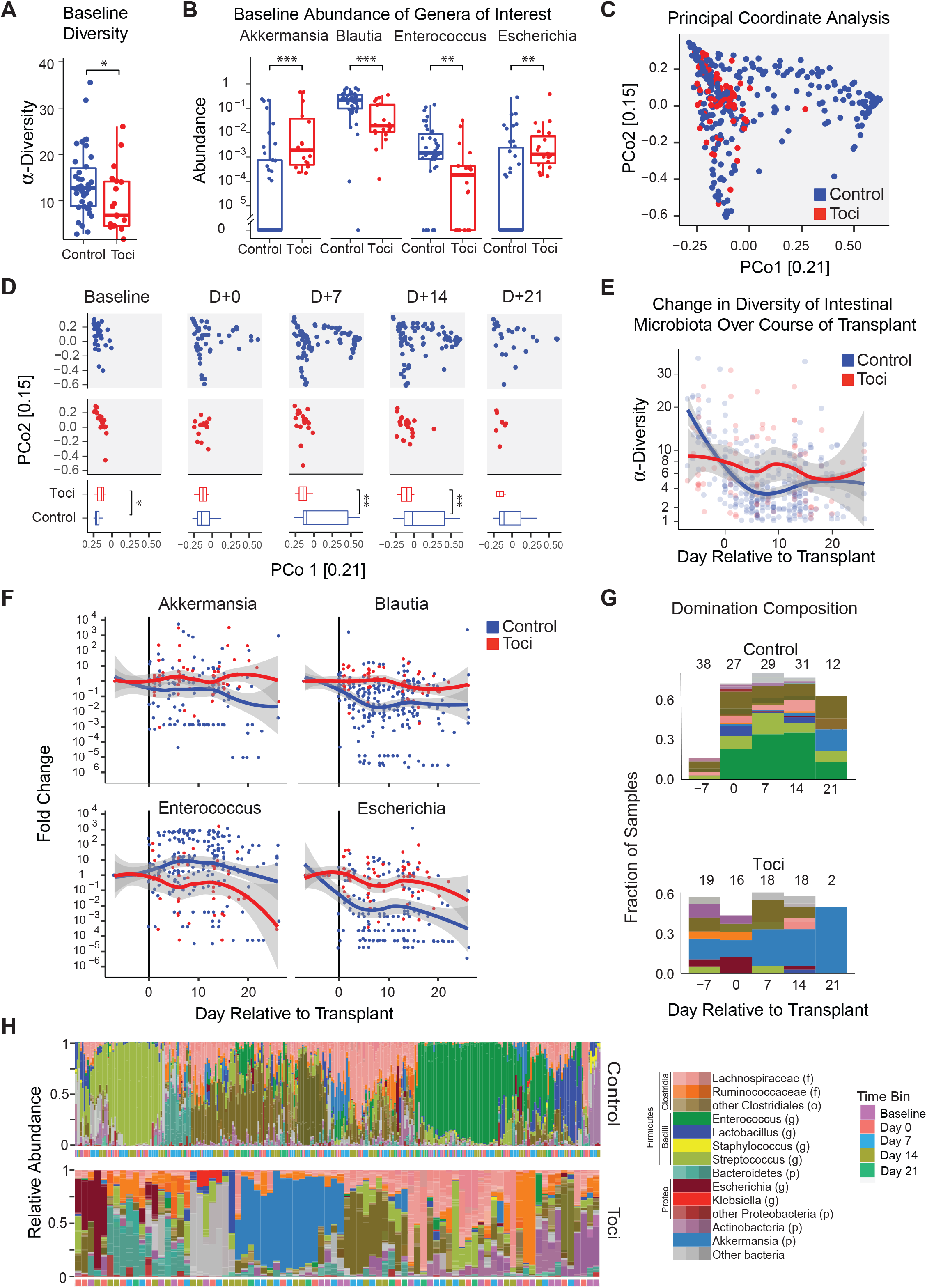
Microbial Diversity and Composition in Tocilizumab and Control Cohorts. (A). Baseline α-diversity, as measured by the inverse Simpson index, in study patients that received toci versus microbiome controls. Baseline samples were collected between day -7 and day -1. (B). Baseline abundance plots of *Akkermansia, Blautia, Enterococcus*, and *Escherichia coli* in these two cohorts. (C). Principal coordinate analysis of Bray-Curtis dissimilarities between samples in which each point is a fecal sample. Samples from toci recipients are largely on the left-hand side of the graph, while samples from the microbiome control are spread throughout the principal-coordinate space. (D). Ordination data from panel C split by specified time windows illustrating the dynamics of fecal compositional shifts along the PC1 axis. In boxplots depicted in panels B and D, the horizontal line in each box represents the median, the lower and upper boundaries of the boxes the interquartile range, and the end of the whisker lines the minimum and maximum values within 1.5 times the interquartile range. (E). Alpha diversity plot over time in tocilizumab recipients compared with microbiome controls. A generalized estimating equation (GEE) was used to compare the α-diversity trends, revealing significant differences between groups both in terms of the trajectory over time (p=0.0005) and the more linear response shape in the toci cohort (p3 =0.001). Each point is a fecal sample; the curves are smoothed averages, and the gray shading are 95% confidence intervals. See Supplementary Table 3 for summary of the GEE model. (F). Fold-change in relative abundances, compared to baseline, of selected genera are plotted and depicted. Each point represents a sample; curves are smoothed averages and shaded areas show 95% confidence intervals. For each fold-change analysis, we considered only patients with detectable abundances of the indicated feature at baseline. (G). Taxa contributing to domination events in samples from the two cohorts. Total stacked-bar height shows the fraction of samples in 7-day bins exhibiting >30% domination by a single amplicon sequence variant (ASV), and the color shows the dominating ASV’s taxonomy according to the color scheme shown below in panel H. (H). Taxonomic composition of all samples is presented as stacked bar graphs and color coded according to the scheme described in Peled *et al*.^11^ Samples are ordered by hierarchical clustering of the beta diversity matrix for each cohort. Panels A, B: Control cohort includes 38 samples from 38 unique patients, Toci cohort includes 19 samples from 19 unique patients. Panels C-H: Control cohort considers 308 samples from 38 unique patients; Toci cohort considers 82 samples from 19 unique patients. Statistics: *p<0.05; **p<0.01; ***p<0.001.

In keeping with this observation, loss of α-diversity in the first two weeks was significantly more rapid in the control cohort compared to toci patients (**Figure 2E**). This was statistically significant as early as day 0 and thereafter at weekly intervals, whereas α-diversity was lower relative to baseline only at three weeks in the toci cohort (**Figure 3A**). Examination of temporal alterations in specific taxonomic groups of interest revealed a significant reduction in *Blauti*a in both cohorts; however, this relative decline occurred at earlier time points in control patients (**Figure 2F** and **Figure 3B**). In addition, *Akkermansia* which had higher baseline abundance in toci patients (**Figure 2A**) further increased by week 3, whereas fold change was unaltered in the control cohort. Most notably, there was a significant increase in the fold change of enterococcal abundance in the control cohort, whereas there was no alteration above baseline in toci patients. In fact, enterococcal domination was observed in approximately a third of control patients but was absent in the toci population (**Figure 2G**, dark green). The differences in domination patterns are illustrated in stacked barplots of taxonomic composition, where frequent dominations by enterococci (dark green) and streptococci (light green) were observed in the control patients; in contrast *Akkermansia* dominations were commonly observed in the toci cohort (**Figures 2G and 2H**, light blue).

**Figure 3:**
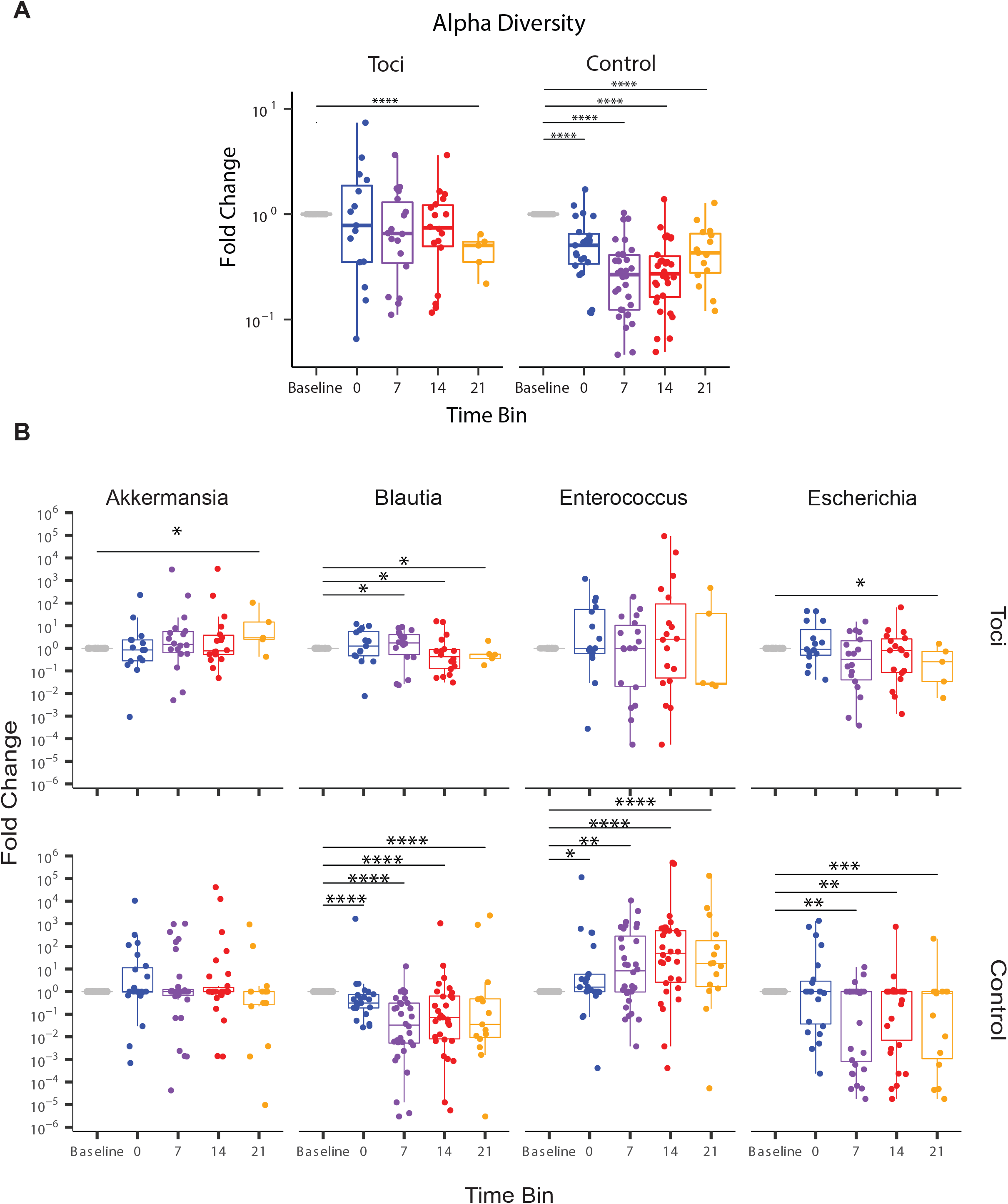
Temporal Alterations in Specific Taxonomic Groups and Alpha Diversity. Fold changes of alpha diversity (panel A) and specified taxa (panel B) are shown over time relative to each patient’s baseline sample. Time is binned by week relative to transplant. For patients with multiple samples within a weekly time window, median abundances were used. Statistics: *p<0.05; **p<0.01; ***p<0.001, ****p<0.0001.

We also explored individual taxa, at the genus level, that exhibited relative expansions or contractions in the two cohorts in multivariable models. These models again highlighted an expansion of Enterococcus in the control cohort that was not observed in the toci cohort (**Supplemental Figure 2**), which we also observed in versions of the models that included terms accounting for time and treatment interactions (**Supplemental Figures 3** and **4**). When antibiotic exposures (**Supplemental Figure 5 and Supplemental Table 2**) were included in the multivariable models, several associations between antibiotic classes and specific general were observed, including expansions of Staphylococcus in carbapenem-exposed samples and reduced abundances of enterococci in samples exposed to the oxazolidone antibiotic linezolid.

## DISCUSSION

In this study, we examined the effect of GVHD prophylaxis with tocilizumab on microbial diversity and composition as well as on overall transplant outcomes. To date, there have been two phase 2^21,32^ and one phase 3^33^ clinical trials that examined the efficacy of tocilizumab for the prevention of GVHD. In all three of these studies, tocilizumab was administered as a single dose the day immediately prior to transplantation followed by standard immune suppression with methotrexate plus either tacrolimus or cyclosporine. Results from the phase 2 studies were consistent in demonstrating a low incidence of both moderate (grades 2-4) and severe (grades 3-4) acute GVHD when compared to historical controls or demographically matched registry data, but with no discernible effect on the incidence of chronic GVHD. In one study,^21^ no cases of lower GI tract GVHD were observed within the first three months, suggesting that tocilizumab mitigated alloreactivity specifically in the GI tract consistent with prior observations in murine models.^19.20^ The phase 3 trial, which was a randomized, placebo-controlled study, demonstrated a reduction in grades 2-4 acute GVHD in tocilizumab-treated recipients, but this did not reach statistical significance. The authors of this trial, however, raised several caveats with respect to the interpretation of the results. Specifically, the investigators identified several potential confounding features, including different patient eligibility criteria when compared to phase 2 trials, a lack of a centralized GVHD grading system in this multi-center trial, a lower-than-expected incidence of acute GVHD in the control arm, and possible underpowering of the trial given that the observed effect size in GVHD-free survival was only 15-20%. Thus, we reasoned that there remain unanswered questions as to the role of tocilizumab in GVHD prevention; particularly with respect to its role in mitigating pathological damage in the GI tract and its impact on the microbiome since this has not been previously assessed.

The design of the current study differed from the three prior reports in several respects. First, we selected a uniform, but high-risk population, that received myeloablative conditioning coupled with peripheral blood stem cell grafts which has been associated with a GRFS incidence that approximates 20%^24^ and is more likely to have low microbial diversity at time of engraftment when compared to reduced intensity or nonmyeloablative transplants.^3^ Secondly, we employed an administration schedule in which patients received a second dose of tocilizumab at day 100. The reason for this was to determine whether additional dosing could reduce the incidence of chronic GVHD since TH17 cells, whose differentiation from naïve CD4^+^ T cells is dependent upon IL-6 signaling,^34^ have been implicated in the pathophysiology of this disorder.^35,36^ In addition, we hypothesized that more sustained protection of the intestinal epithelium from immune-mediated attack by IL-6 blockade could further promote the production of short chain fatty acids which have been shown to correlate with protection from chronic GVHD.^37^ Another reason for this schedule was to ascertain whether extended administration of tocilizumab could prevent late acute GVHD where GI tract disease is the primary driver of morbidity, as this had been reported in a prior study.^21^

A main goal of the trial was to determine the probability of GRFS which was calculated to be 38% at one year post transplantation. This was statistically significant when compared to the pre-specified historical control value of 20%, indicating that the primary endpoint of the study was met. Analysis of secondary endpoints revealed a low incidence of grades 3-4 acute GVHD which was similar to that reported in other phase 2 studies.^21,32^ Triggering of GRFS was primarily due to the development of chronic GVHD that required systemic immunosuppression, indicating that the additional dose of tocilizumab had no apparent effect on preventing this disease. Despite the observed incidence of chronic GVHD, however, severe disease occurred in only two patients. Consequently, TRM in tocilizumab-treated patients was low with only one death attributable to chronic GVHD, and no acute GVHD related-mortality. The remaining three deaths in the trial were due to disease recurrence, resulting in an overall probability of disease-free survival of 86% at one year.

Importantly, only one of 29 patients developed lower GI tract GVHD in the first 100 days and there were no further cases thereafter within the first-year post transplantation, providing evidence that IL-6 inhibition suppresses inflammation in the GI tract. We hypothesized that attenuation of intestinal inflammation by IL-6 blockade^19,20^ would mitigate the microbiome injury that accompanies allogeneic HSCT. Although the control cohort was assembled *post hoc* from patients at another center, transplant characteristics were well-matched with trial participants, and samples from both centers were centrally sequenced and computationally analyzed. Even though baseline diversity was higher in the control cohort, the loss of diversity was significantly attenuated in tocilizumab-treated patients. The dramatic shifts in global microbiome composition that have been well described in allogeneic HSCT recipients^3,11^ were observed in the control cohort, but to a markedly lesser extent in tocilizumab recipients. Patients who go on to develop GVHD frequently have a shift in their microbiota from Clostridiales to Lactobacillales/Enterobacteriales. A consistent observation has been the expansion of enterococcus which is associated with the emergence of vancomycin-resistant strains and the subsequent development of bacteremia which can lead to increased mortality.^7,9^ Loss of microbial diversity has been shown to be more pronounced in patients with GI tract GVHD, and expansion of enterococcus is particularly prominent in this patient population.^8^ Notably, this pattern of enterococcal domination has been highly consistent and reproducible across transplant centers and geographical locations, even across different patterns of antibiotic exposure.^11,31^ In tocilizumab-treated patients, we saw little evidence of enterococcal dominance and only one case of enterococcal bacteremia. This contrasted with cohort-matched patients from MSKCC where enterococcal domination occurred in approximately 30% of recipients which was consistent with previous incidence results obtained from a much larger patient population at this same institution.^11^

An interesting observation was the relative abundance of *Akkermansia muciniphilia* that was present in a significant number of patients that were treated with tocilizumab. This organism is a gram-negative bacterium that inhabits the large intestine where it accounts for 1-4% of intestinal bacteria in normal adults.^38^ *Akkermansia muciniphilia* is a mucus-degrading bacterium that resides in the mucus layer next to intestinal epithelia cells^39^ and whose abundance has been inversely correlated with a variety of diseases, including obesity, diabetes, and metabolic syndrome.^40^ Conversely, dietary supplementation of *Akkermansia muciniphilia* appears to improve parameters of insulin resistance and augment weight loss in humans.^41^ Mechanistically, this organism has been shown to enhance gut barrier function by increasing the number of Paneth and goblet cells, the production of antimicrobial metabolites, and the proliferation of intestinal stem cells,^42,43^ suggesting a salutary role in preserving intestinal barrier function, although this has not been observed in all studies.^31,44^ The emergence of *Akkermansia* observed only after tocilizumab exposure raises the possibility that IL-6 blockade may have preserved the production of mucus, on which this organism thrives, by colonic goblet cells that are otherwise damaged by conditioning and alloreactivity.^45^

In summary, this study demonstrates that an extended course of tocilizumab administration is effective for the prevention of lower GI tract GVHD in patients receiving myeloablative conditioning followed by transplantation with allogeneic peripheral blood stem cell grafts. In addition, loss of microbial diversity and enterococcal domination within the GI tract is attenuated in tocilizumab-treated recipients. Thus, GVHD prophylaxis with adjunctive tocilizumab represents a potential therapeutic approach for remodeling the intestinal microbiome and preventing the emergence of potentially pathogenic organisms.

## Data Availability

All data produced in the present study are available upon reasonable request to the authors

## AUTHOR CONTRIBUTIONS

S.C. designed the study, enrolled patients on the trial, provided clinical care of patients in the study, analyzed data, and wrote the paper. A.S. and D.E. provided statistical analysis. K.M. and L.S. assisted in the acquisition of data. S.A., W.L., P.N.H., M.H., N.N.S., L.R., and J.H.J. enrolled patients on the trial, provided clinical care of patients, and edited the paper. A.C., N.W., G.A., T.F., J.U.P. and M.vdB performed microbiome computational analyses and edited the paper. W.R.D. designed the study, provided clinical care, analyzed data, and wrote the paper.

## ACKNOWLEDGMENTS

This research was supported by funding from K08 HL143189 (J.U.P.), and the MSKCC Cancer Center Core Grant NCI P30 CA008748 (J.U.P.). This research was also supported by the Parker Institute for Cancer Immunotherapy at Memorial Sloan Kettering Cancer Center; J.U.P. is a member of the Parker Institute for Cancer Immunotherapy.

