## Supplemental Data for "Mitigation of gastrointestinal graft versus host disease with tocilizumab prophylaxis is accompanied by preservation of microbial diversity and attenuation of enterococcal domination"

### SUPPLEMENTAL METHODS

**Patient Population.** Patients were excluded if they had had a prior allogeneic HSCT, Karnofsky Performance Score <60%, uncontrolled bacterial, viral or fungal infections at time of enrollment, active hepatitis B or C virus infection, or known human immunodeficiency virus (HIV) positive, active CNS involvement with malignancy, receiving cord blood or haploidentical allograft, prior intolerance or allergy to tocilizumab, use of rituximab, alemtuzumab, ATG or other monoclonal antibody at time of conditioning regimen, or history of diverticulitis, Crohn’s disease, ulcerative colitis or a demyelinating disorder, or any current uncontrolled cardiovascular conditions, including uncontrolled ventricular arrhythmias, NYHA class III or IV congestive heart failure, uncontrolled angina, or electrocardiographic evidence of active ischemia or active conduction system abnormalities.

**Conditioning Regimens and GVHD Prophylaxis.** Contraindications to receipt of day 100 Tocilizumab dose were as follows: (1) Grade 4 (according to CTCAE version 5 hematologic or nonhematologic events. Dosing was allowed upon recovery to grade ≤3. Transfusion to increase platelet and hemoglobin was allowed. Grade 3 febrile neutropenia and grade 3 increased AST/ALT/bilirubin were also contraindications to dosing of tocilizumab; recovery to grade 2 or lower was allowed for tocilizumab dosing within the time frame of day +86 through day +114, (2) Disease relapse or progression of the primary malignancy for which treatment was indicated, (3) Donor leukocyte infusion prior to planned second dose of tocilizumab for any indication: infection, disease control or to address mixed chimerism, (4) Grade III–IV acute GVHD. Patients with grade I–II acute GVHD and on topical or systemic corticosteroids therapy (equivalent to prednisone ≤20 mg/d) could receive a second dose of tocilizumab, and (5) Active chronic GVHD requiring systemic therapy, and (6) Acute GVHD treated with tocilizumab before day +100.

**Infection Prophylaxis.** At MCW, anti-bacterial prophylaxis was with levofloxacin with onset of neutropenia, followed by first-line therapy with cefepime for first fever. At MSKCC, patients were treated with ciprofloxacin at time of neutropenia and then switched, most commonly, to piperacillin/tazobactam with onset of first fever. Empiric antibiotics were continued until engraftment at both institutions.

**Microbiome Sequencing**. Microbiome profiles of fecal samples were generated as described previously.[11] Briefly, bacterial cells were disrupted by silica bead-beating in phenol:chloroform followed by ethanol-based isolation of nucleic acids and amplification of the 16S ribosomal-RNA gene V4-V5 variable region on the Illumina MiSeq platform. Amplicon sequence variants (ASVs) were called using the DADA2 algorithm and mapped in the NCBI 16S rRNA sequence database using BLAST.

**Statistical Analysis of Clinical Outcomes.** Demographic and other baseline data, such as disease characteristics, as well as outcome measures, are presented for patients. Categorical data, such as gender, etc., are presented by frequencies and percentages. Descriptive summary statistics (e.g., frequency, mean, median, range and standard deviation) are used to present numeric data. Time-to-event outcomes with and without competing risks were analyzed using Kaplan-Meier and Nelson-Aalen estimates, respectively, and presented with 95% confidence intervals computed with the complimentary log-log transformation. All follow up was administratively censored at 1 years. Binary outcomes were analyzed using proportions with 95% confidence intervals. GRFS was estimated using the Kaplan-Meier estimator and plotted with a 95% confidence band. An event for this outcome is defined as grade III-IV acute GVHD, chronic GVHD requiring systemic therapy, relapse, or death. Patients who are alive without GVHD were censored at the last follow-up. The 12-month GRFS was compared to the prespecified historical control value of 20% using a one-sided *z*-test. Clinical outcome analyses were performed using SAS 9.4 (SAS Institute, Cary, NC).

**Statistical Analysis of Microbiome Comparisons.** Wilcoxon tests were used to compare groups in Figures 2A, 2B, and 2D. P-values were corrected for multiple comparisons using the Benjamini-Hochberg method.

**GEE Model**

A Generalized Estimating Equation model was generated using the R package *geepack* [46-48] to associate treatment group/center and time with alpha diversity, grouping by patient. Polynomial time was included to account for the non-linear nature of the alpha diversity profiles over time. A model summary for the GEE analysis is depicted in **Supplemental Table 3**.

**Ordination**

PCoA was performed on the Bray-Curtis distances of the genus-level counts using the R package *ape* package.[49] This PCoA was split out by 7-day bins in Figure 2D to highlight the dynamics over time as they differed across treatment groups/centers.

**Multivariate analysis**

Data for multivariate analyses were encoded as follows: ages were binned into three groups based on clinical decision-making ("39 and under", "40-59", and "60 or over"), and disease was grouped into either "acute leukemia" or "other".

Maaslin2[50] was used to investigate multivariate relationships between taxonomic features and patient characteristics. MaAsLin2 was run with a minimum relative abundance threshold 2x10^-3 (corresponding to the limit of detection given the inclusion criteria of a minimum 1000 reads per sample), and a minimum prevalence of 5%, or 19 samples. The base linear model was used on the log-scaled abundances. Patient ID was included as a random effect. Hits were considered significant if they had a Benjamini-Hochberg corrected p-value less than 0.05.

Two models were considered. First, a bivariate model of time and treatment (toci vs control) was constructed to show taxa associated with the differences in alpha/beta diversity expanding beyond the select taxa shown in Figure 2F and Figure 3 **(see Supplemental Figures 2-4)**. A second model was run that included drug exposures, patient characteristics, and disease. Drug groups are described in Supplemental Table 1; in short samples were considered “exposed” to a group of antibiotics if the patient had received any drug within that group as recently as the previous calendar day (**Supplemental Figure 5**).

#### SUPPLEMENTAL FIGURE LEGENDS

Supplemental Figure 1: **Engraftment.** (A). Cumulative incidence of patients that achieved an absolute neutrophil count (ANC) >500/mm3 for three consecutive days. (B). Cumulative incidence of patients that achieved an unsupported platelet count >20,000/$mm^3$.

Supplemental Figure 2: **Bivariate Analysis.** Taxa associated with changes over time and/or by treatment group/center. The genus-level abundances were analyzed by MaAslin2[28] using day relative to transplant and group (tocilizumab participants vs microbiome controls) as fixed effects, and patient ID as a random effect. Taxonomic changes over time are shown in the day relative to transplant (drt) column, with red indicating a relative increase and blue indicating a relative decrease. Taxa differentially abundant by treatment group/center are shown relative to the tocilizumab participants in the column labeled "MSK". Benjamini-Hochberg corrected p-values are shown as shading, and the size represents the absolute value of the magnitude of the change. In a few cases where the taxa could not be assigned to a genus, they were classified at the family level (e.g., f_Lachnospiraceae).

Supplemental Figure 3: **Bivariate Analysis with Interaction Term.** The previous model shown in Supplemental Figure 2 was modified to include a time and treatment interaction term.

Supplemental Figure 4: **Bivariate Analysis with Interaction Term and Marginal Effects.** Coefficients are shown, as in Supplemental Figure 3, along with the calculated marginal coefficients representing the change in time relative to the MSK/control group for those taxa with significant interactions (no p-value is available for the computed marginal effect column).

Supplemental Figure 5: **Multivariate Analysis.** The model shown in Supplemental Figure 2 was modified to include drug-group exposure and selected patient characteristics. The drug groups are described in

Supplemental Table 2. Disease was categorized into either “Acute leukemia” or “other” for the purposes of modeling, taxonomic changes relative to the “Acute leukemia” group are shown; age is shown relative to the “40-59” group.

### Supplemental Figure 1

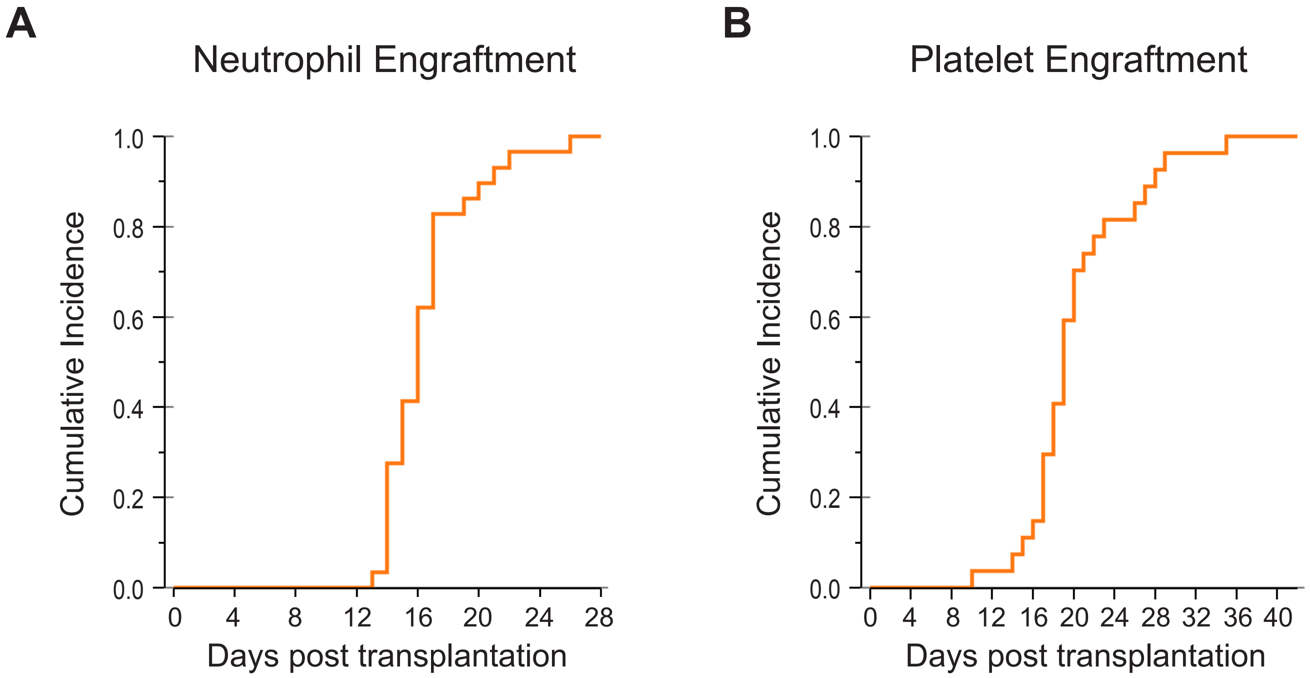

### Supplemental Figure 2

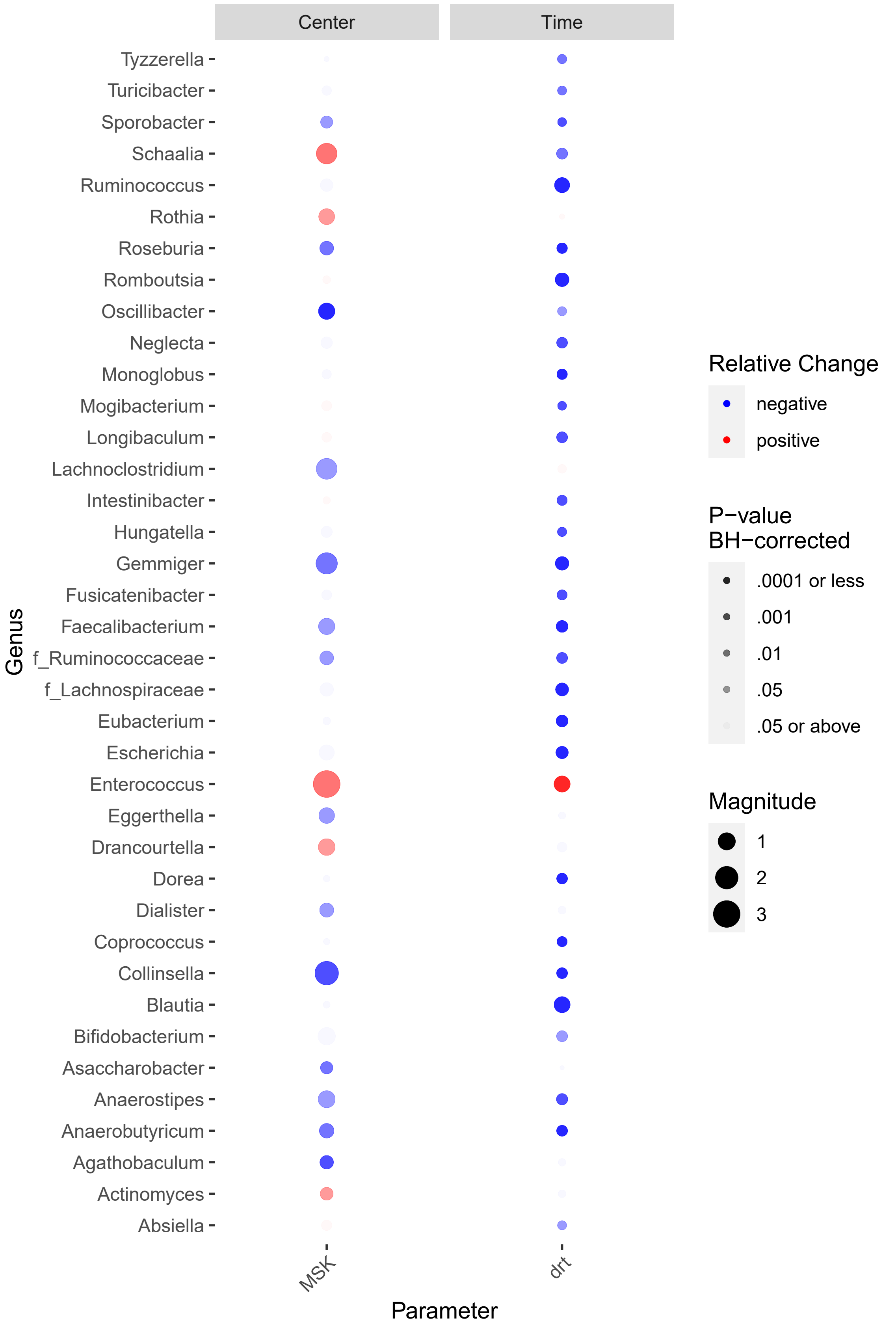

### Supplemental Figure 3

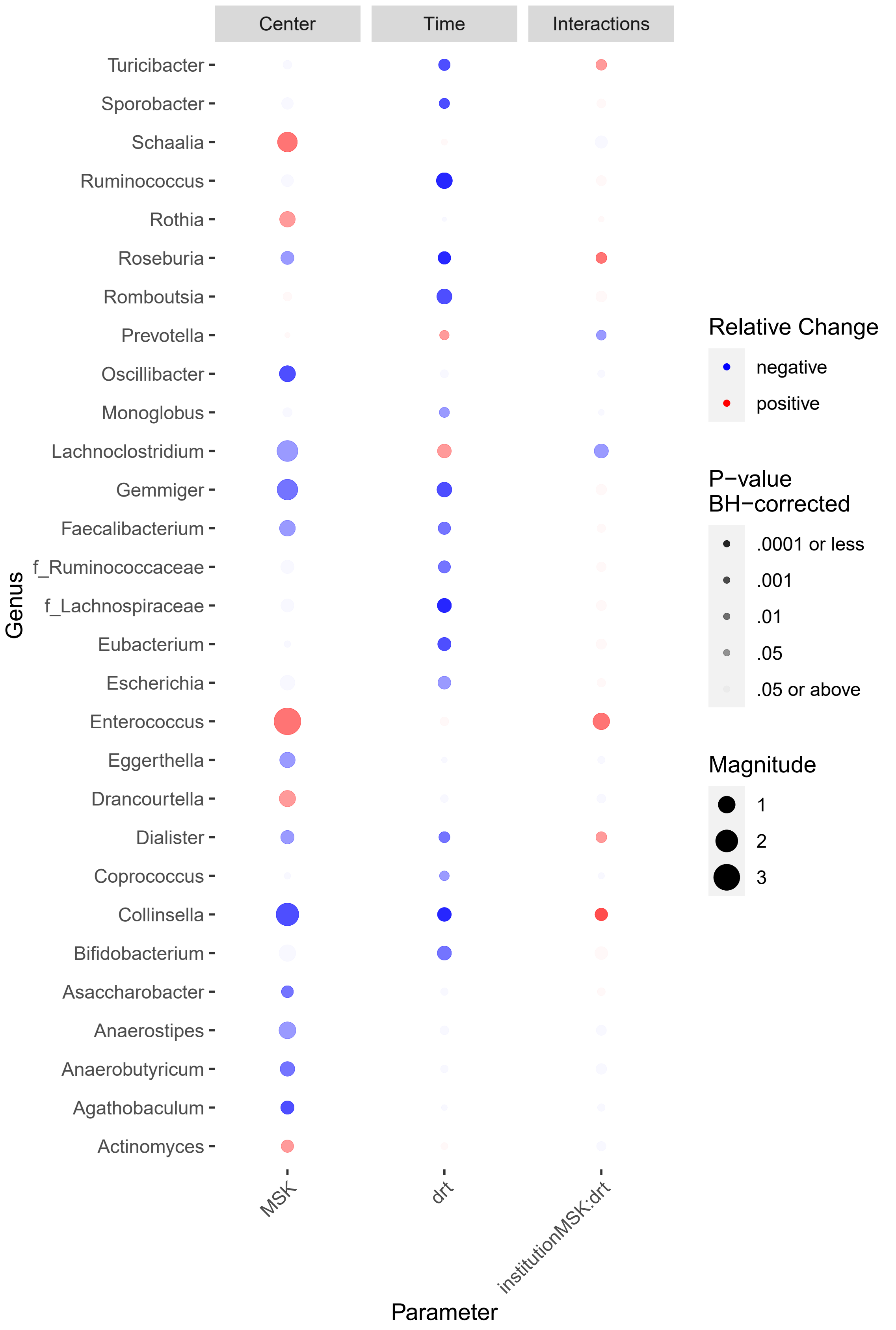

### Supplemental Figure 4

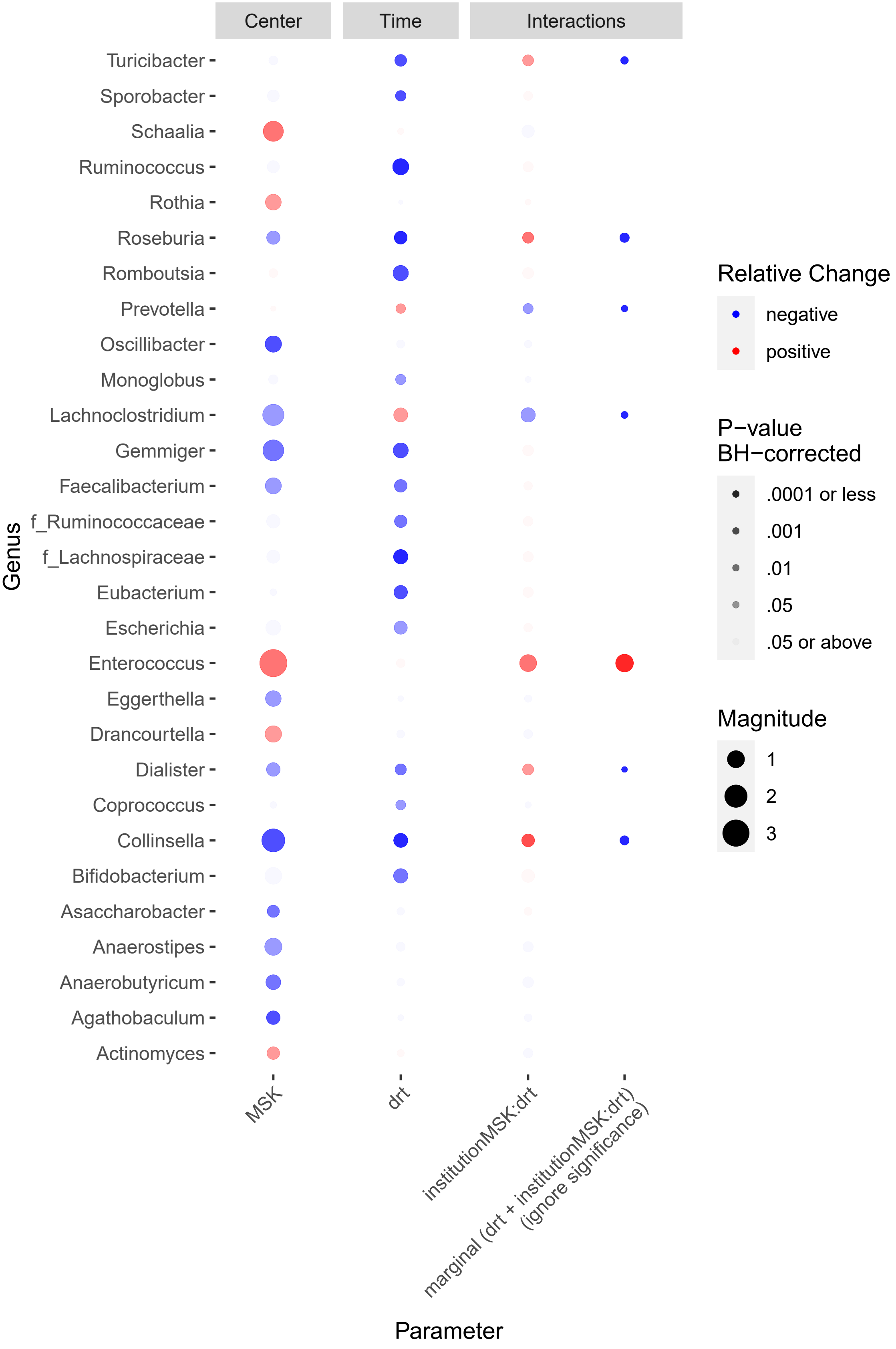

### Supplemental Figure 5

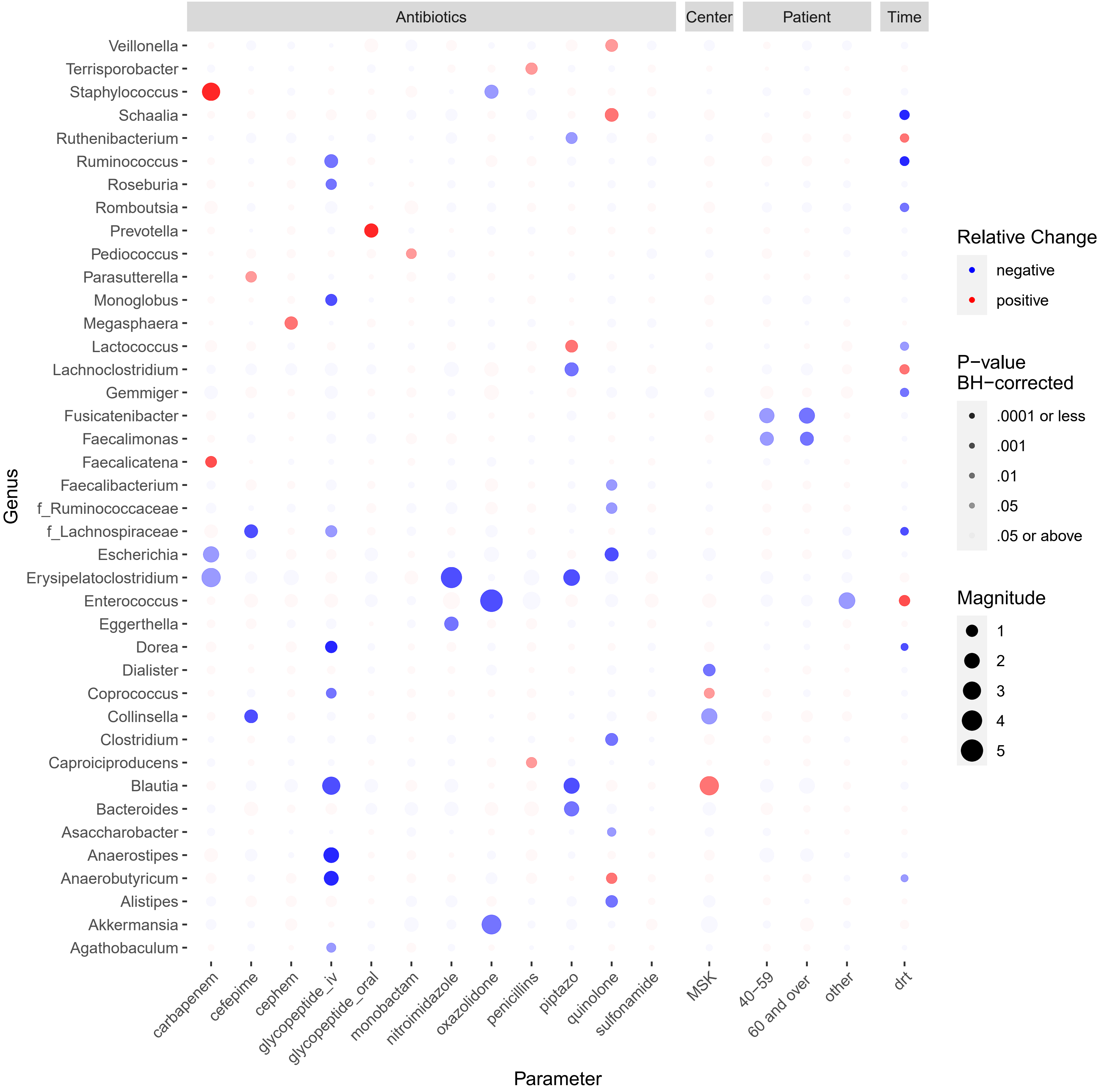

**SUPPLEMENTAL TABLE 1. DEFINITION OF EVENTS AND COMPETING RISKS FOR SPECIFIC OUTCOMES**

| Outcome | Event(s) | Competing risks |
|---|---|---|
| **Event-free survival** | | |
| GRFS | Grade 3-4 aGVHD, Relapse, Death or cGVHD requiring systemic therapy | None |
| Disease-free survival | Relapse or death | None |
| Overall survival | Death | None |
| **Cumulative incidence** | | |
| Treatment related mortality | Death | Relapse |
| Relapse | Relapse | Death w/o relapse |
| Acute GVHD | Grade II-IV aGVHD | Relapse or death |
| Chronic GVHD | Chronic GVHD (Mild-Severe) | Relapse or death |
| Neutrophil engraftment | $1^{st}$ of 3 consecutive days with an ANC>500 | Death w/o engraftment |
| Platelet engraftment | $1^{st}$ day of a sustained platelet count above 20,000 w/o any platelet transfusions for the preceding 7 days | Death w/o engraftment |

All outcomes used loss to follow up / end of study as censoring.

#### SUPPLEMENTAL TABLE 2. ANTIBIOTIC EXPOSURE

| Antibiotic | Group | Toci | Control |
|---|---|---|---|
| vancomycin(iv) | glycopeptide(iv) | 2 | 37 |
| ciprofloxacin | quinolone** | 7 | 34 |
| piperacillin/tazobactam | piptazo** | 5 | 19 |
| sulfamethoxazole/trimethoprim | sulfonamide** | 5 | 6 |
| cefepime | cefepime** | 6 | 5 |
| cefazolin | cephem** | - | 5 |
| metronidazole | nitroimidazole** | 3 | 4 |
| levofloxacin | quinolone** | 28 | 3 |
| vancomycin(oral) | Glycopeptide (oral) | 3 | 3 |
| aztreonam | monobactam** | 1 | 3 |
| penicillin | penicillins** | - | 3 |
| ceftriaxone | cephem** | 2 | 2 |
| linezolid | oxazolidone** | 2 | 2 |
| cefuroxime | cephem** | 1 | 2 |
| meropenem | carbapenem** | 1 | 2 |
| ampicillin | penicillins** | - | 1 |
| azithromycin | macrolide | - | 1 |
| ceftazidime | cephem** | - | 1 |
| cephalexin | cephem** | - | 1 |
| imipenem/cilastatin | carbapenem** | - | 1 |
| amoxicillin/clavulanate | penicillins** | 2 | - |
| dapsone | antibacterial (other) | 1 | - |

The number of patients in each cohort exposed to different antibacterial antibiotic drugs is tabulated. 'Group' is a grouping of different antibiotics defined for this multivariable analysis based broadly on pharmacologic classification with some customization, for example intravenous and oral vancomycin are grouped separately. Only drug groups administered in at least one member of each cohort were considered. For the multivariate model, exposure was encoded as binary where a sample was considered 'exposed' if the patient had received a dose of any drug from the group, considering a window extending back to day-30 relative to transplant. Asterisks indicate this drug group was included in the multivariate model in Supplemental Figure 6.

**SUPPLEMENTAL TABLE 3. MODEL SUMMARY FOR GEE ANALYSIS (Accompanies Figure 2E)**

| term | estimate | std. error | statistic | P value |
|---|---|---|---|---|
| Intercept | 8.61 | 0.498 | 299.17 | 0.000000 |
| Center(MCW) | 0.55 | 1.125 | 0.24 | 0.626647 |
| Day | -0.82 | 0.099 | 68.12 | 0.000000 |
| Day^2 | 0.03 | 0.005 | 51.78 | 0.000000 |
| Center: Day | 0.66 | 0.189 | 12.16 | 0.000488 |
| Center: Day^2 | -0.03 | 0.010 | 10.54 | 0.001167 |
